# Effectiveness of electroacupuncture in the treatment of non-alcoholic fatty liver disease : a systematic review and meta-analysis of randomized controlled trials

**DOI:** 10.1101/2025.02.09.25321958

**Authors:** Xiaoguang Lin, Shuo Niu, Jiongliang Zhang, Yuting Wang, Luwen Zhu

## Abstract

**Objectives:** Nonalcoholic Fatty Liver Disease (NAFLD) is a metabolic disease syndrome with progressive disease development leading to cirrhosis and liver cancer. Electroacupuncture (EA) is a non-pharmacological therapy. The aim of this systematic review is to assess the efficacy and safety of electroacupuncture in the treatment of NAFLD.

**Design:** Systematic review and meta-analysis.

**Data sources:** A comprehensive database search was conducted using PubMed, Embase, the Cochrane Library, Web of Science, Cnki, Wan Fang, VIP to identify randomized controlled trials (RCTs) investigating the impact of electroacupuncture (EA) on Non-alcoholic fatty liver disease. RCTs published until July 31, 2024, that met our predetermined inclusion and exclusion criteria were included. Data extraction, literature review, and assessment of the methodological quality of the trials were performed. The meta-analysis was conducted using StataSE version 16.

**Eligibility criteria for selecting studies:** We included randomized controlled trials (RCTs) about electroacupuncture for non-alcoholic fatty liver disease. The primary indicators :1. Alanine Aminotransferase (ALT), 2. Aspartate Aminotransferase (AST), 3. Gamma-Glutamyl Transpeptidase (GGT), 4. Triglycerides (TG), 5. Total Cholesterol (TC). Secondary indicators:1. PCIII (Pre-type III collagen), 2. HA (hyaluronidase), 3. LN (laminin), 4. CIV (collagen type IV).

**Data extraction and synthesis:** Two independent reviewers conducted separate searches of the databases, assessed eligible articles for inclusion and employed the Cochrane Collaboration’s tool for assessing the risk of bias. The analyses were performed using RevMan 5.3 and StataSE version 16 software. The mean difference (MD) with 95% CI was employed to analyze continuous outcomes.

**Results:** The primary outcome was Triglycerides (TG), Total cholesterol (TC), Alanine aminotransferase (ALT), Aspartate aminotransferase (AST), Gamma-glutamyl transpeptidase (GGT). The secondary outcomes were Laminin (LN), Hyaluronidase (HA), Pre-type III collagen (PCIII), Collagen type IV (CIV). Our meta-analysis includes 986 patients from 11 RCTs incorporated. Main result : The Triglycerides (TG) (SMD=-0.84, 95%CI=-1.03,-0.65, P=0.000), Total cholesterol (TC) (SMD=-0.48, 95%CI=-0.62,-0.35, P=0.000), Alanine aminotransferase (ALT) (SMD=-1.48, 95%CI=-1.98,-0.98, P=0.000), Aspartate aminotransferase (AST) (SMD=-0.84, 95%CI=-1.08, −0.60, P=0.000), Gamma-glutamyl transpeptidase (GGT) (SMD=-1.21, 95%CI=-1.59, −0.84, P=0.000). There was a statistically significant difference between the electroacupuncture group and the drug group in terms of improvement in liver blood indices such as lipids. The secondary Laminin (LN) (SMD=-1.90, 95%CI=-2.13, −1.67, P=0.000), Hyaluronidase (HA) (SMD=-3.42, 95%CI=-3.72, −3.12, P=0.000), Pre-type III collagen (PCIII) (SMD=-0.41, 95%CI=-0.62, −0.19, P=0.000), Collagen type IV (CIV) (SMD=-1.08, 95%CI=-1.28, −0.87, P=0.000). There was a statistically significant difference between the electroacupuncture group, and the drug group compared to the drug group in terms of the four indices of liver fibrosis.

**Conclusions:** Compared to the control group, the addition of electroacupuncture significantly improved Triglycerides (TG), Total cholesterol (TC), Alanine aminotransferase (ALT), Aspartate aminotransferase (AST), Gamma-glutamyl transpeptidase (GGT), Laminin (LN), Hyaluronidase (HA), Pre-type III collagen (PCIII), Collagen type IV (CIV)with laboratory blood indicators of Non-alcoholic fatty liver disease. Electroacupuncture helps improve lipid levels and fibrosis progression in non-alcoholic fatty liver disease.

**PROSPERO registration number:** CRD42024626611

**STRENGTHS AND LIMITATIONS OF THIS STUDY:**

- The search strategy was robust and thorough, and strict filtering criteria were implemented.
- Subgroup analyses based on different interventions were conducted to strengthen the reliability of the findings.
- The quality of the included literature was not sufficiently high and did not involve blinding.

## Introduction

Non-alcoholic fatty liver disease (NAFLD) encompasses a spectrum of liver conditions, ranging from simple fatty liver to non-alcoholic steatohepatitis (NASH). It is associated with visceral adiposity and metabolic syndrome, and its prevalence is as high as that of type 2 diabetes [1]. Approximately 5% of patients with non-alcoholic fatty liver disease (NAFLD) may progress to cirrhosis or hepatocellular carcinoma, conditions that have become significant indications for liver transplantation [2]. It is a chronic non-communicable disease characterized by multi-systemic involvement and is causally linked to obesity, metabolic syndrome, and type 2 diabetes mellitus. These conditions collectively contribute to the development of cirrhosis, cardiovascular and cerebrovascular diseases, chronic kidney disease, as well as intra- and extra-hepatic malignancies. Among these, atherosclerotic cardiovascular and cerebrovascular diseases, as well as malignant tumors, are the primary causes of disability and death in patients with non-alcoholic fatty liver disease (NAFLD) [3]. Non-alcoholic fatty liver disease (NAFLD) is not merely a standalone health issue; it can be complicated by a range of serious conditions, including cardiovascular disease, type 2 diabetes mellitus, kidney disease, and liver cancer. These complications can significantly affect a patient’s quality of life and life expectancy. NAFLD has emerged as a major global public health concern, imposing a substantial clinical and economic burden that demands our immediate attention.

Epidemiological surveys indicate that, as of May 2021, the global prevalence of non-alcoholic fatty liver disease (NAFLD) is 32.4 percent, according to a meta-analysis. This condition is significantly more prevalent in men (39.7 percent) than in women (25.6 percent) [4]. According to estimates, by 2040, over half of the adult population is projected to have non-alcoholic fatty liver disease (NAFLD). The most significant increases are anticipated among women, smokers, and individuals without metabolic syndrome [5]. The prevalence of non-alcoholic fatty liver disease (NAFLD) varies by race and region, with notably higher rates observed in Latin America and the Middle East and North Africa compared to other parts of the world [6]. Environmentally, airborne particulate matter is believed to increase the risk of non-alcoholic fatty liver disease (NAFLD), exacerbate metabolic disorders and inflammatory dysfunction in patients with NAFLD, and promote the progression of NAFLD to non-alcoholic steatohepatitis (NASH). Exposure to environmental factors and pollutants significantly contributes to the elevated risk of developing NAFLD, as these factors can affect liver health and metabolic function through various mechanisms [6].

A variety of lipid-lowering and hepatoprotective drugs, including silymarin (bin), polyene phosphatidylcholine, ursodeoxycholic acid, and statins, are currently used in regimens for the treatment of non-alcoholic fatty liver disease (NAFLD). However, it typically takes weeks or even months for these medications to exhibit lipid-lowering and liver-protective effects [7,8]. These oral medications have various contraindications and side effects. Therefore, safer and more effective treatments are still needed to slow disease progression in patients with non-alcoholic fatty liver disease (NAFLD), lower blood lipid levels, and protect the liver. As a traditional Chinese alternative healthcare approach, acupuncture is gaining increasing attention and recognition both in China and internationally. Acupuncture is widely recognized as a safe and effective alternative therapy in clinical practice. Numerous clinical studies have demonstrated that acupuncture can alleviate obesity [11] and hypertension [12]. One of the groundbreaking advancements in acupuncture is the development of electroacupuncture (EA). EA involves the insertion of needles into the skin or deep muscles, followed by electrical stimulation. Compared to manual acupuncture, electroacupuncture can yield similar or even superior results. It has been shown to be more effective than manual acupuncture in reducing pain and enhancing grip strength. Additionally, electroacupuncture offers continuous stimulation for an extended duration, which may result in further therapeutic benefits [13].

The pathogenesis of non-alcoholic fatty liver disease (NAFLD) is primarily attributed to the accumulation of fat in the liver, which is typically associated with various metabolic abnormalities. Some experiments have demonstrated that acupuncture may exert a lipolytic effect by enhancing sympathetic nerve function and increasing the levels of adrenaline and norepinephrine in the bloodstream. This process activates adenylate cyclase on the cell membrane, leading to an increase in intracellular cyclic adenosine monophosphate (cAMP), which ultimately results in a lipolytic effect and contributes to the reduction of lipid levels [14,15]. In contrast, insulin resistance (IR), a critical factor in the pathogenesis of non-alcoholic fatty liver disease (NAFLD), leads to increase de novo lipid synthesis, promotes the transport of fatty acids to the liver, and inhibits the β-oxidation of free fatty acids (FFA), thereby further contributing to the accumulation of hepatic lipids. One study has demonstrated that acupuncture reduces body mass index (BMI) and waist circumference while improving insulin resistance [16, 17]. An animal study found that electroacupuncture regulates autophagy through the hepatic SIRT1/ATG7 pathway, which is one of the mechanisms by which it improves insulin resistance (IR). This provides a strong theoretical basis for the use of electroacupuncture in the prevention and treatment of IR and its related diseases [18].

Although there are no established guidelines, electroacupuncture has been utilized for the treatment of patients with non-alcoholic fatty liver disease (NAFLD) [19,20]. To date, no meta-analyses have been conducted specifically addressing the use of electroacupuncture for NAFLD. Therefore, the objective of this systematic review is to evaluate the effectiveness of electroacupuncture in treating NAFLD and to offer practical guidance for clinical practice.

## Methods and materials

We have pre-registered our review protocol (https://www.crd.york.ac.uk/prospero/) under registration number CRD42024626611. All reviewers receive the same training to ensure integrity and consistency throughout the review process. We developed methods based on the Preferred Reporting Items for Systematic Reviews and Meta-Analyses (PRISMA) criteria and meta-analysis protocols and the Cochrane Handbook for the Evaluation of Systems of Intervention to ensure the accuracy of these systematic reviews and meta-analysis.

### Diagnostic criteria

Patients included in the study were required to be diagnosed based on the established diagnostic criteria outlined in the for the Prevention and Treatment of Non-Alcoholic Fatty Liver Disease: 2018 Update, published by the Fatty Liver and Alcoholic Liver Disease Group of the Hepatology Branch of the Chinese Medical Association [21].

### Inclusion and exclusion criteria

#### The inclusion criteria

1. The study was conducted on individuals with confirmed diagnosis of non-alcoholic fatty liver disease (NAFLD). (2) The study design is a strictly limited RCT. (3) The experimental group received electroacupuncture, either alone or in combination with other methods. The control group administered a drug. (4) The language was limited to English or Chinese.

The exclusion criteria:

1. Animal studies, case reports, self-controls, non-randomized controlled trials, class randomized controlled trials, and duplicate published studies. (2) Articles with incomplete data. (3) The patient clearly has several significant comorbidities, including cancer and infectious diseases.

### Search strategy

We conducted electronic and manual searches of seven databases for all randomized controlled trials of electroacupuncture in NAFLD from the start of the database to 31 July 2024 by searching the databases. These databases included PubMed, Web of Science, Embase, Cochrane Library, China Knowledge Network (CNKI), Chinese Scientific Journal Database (VIP), and Wanfang. Use medical subject terms and keywords to search for articles. (NAFLD, electroacupuncture, randomized controlled trials) combined with Boolean logic operators. Electronic searches included the use of subsequent search terms: (NAFLD [Mesh Terms] or NAFLD [Title/ Abstract] or Fatty Liver Disease [Title/Abstract] and (Electroacupuncture [Mesh Terms] or Electroacupuncture Therapy [Title/Abstract] or Electroacupuncture [Title/Abstract]) and (randomized controlled trials [Title/Abstract]). We adapted the search terms of the different databases to suit their search criteria.

## Data collection and analysis

### Study selection

Two reviewers (XGL and SN) independently assessed the screening results. The initial review involved evaluating titles, abstracts, and keywords. This was followed by a comprehensive examination of the full texts of potential studies that met the inclusion criteria. If any disagreements arose during this process, they were resolved through negotiation with a third-party reviewer (JLZ).

### Data extraction and management

The following information will be obtained by our researchers (YTW and JLZ) from the included RCTs, respectively, independently using pre-designed extraction scales: first author, date of publication, sample size, age, intervention, acupoints, frequency, treatment duration, outcome, efficacy, and adverse effects. We will endeavor to contact the authors if any information is missing or unclear.

### Outcome measures

The primary indicators for Non-Alcoholic Fatty Liver Disease (NAFLD) include several laboratory tests used to diagnose NAFLD and assess hepatocellular injury:

1. Alanine Aminotransferase (ALT): ALT is the most utilized sensitive marker for monitoring liver function and screening for liver disease, particularly in cases of hepatocellular injury, where ALT levels are elevated. Elevated ALT levels serve as a crucial marker in the diagnosis of NAFLD. 2. Aspartate Aminotransferase (AST): AST, in conjunction with ALT, is a sensitive indicator of hepatocellular injury. These two enzymes are often measured together, and the AST/ALT ratio holds clinical significance. 3. Gamma-Glutamyl Transpeptidase (GGT): GGT levels are frequently elevated in patients with NAFLD and are associated with advanced fibrosis and increased mortality risk. 4. Triglycerides (TG): TG is a lipid component linked to the development of NAFLD, particularly in the context of insulin resistance and metabolic syndrome. 5. Total Cholesterol (TC): The relationship between TC and NAFLD has been explored, especially when evaluated alongside other lipid markers such as high-density lipoprotein (HDL) and low-density lipoprotein (LDL).

### Secondary indicators

PCIII, HA, LN and CIV are the indicators of four tests for liver fibrosis, which represent:

1. PCIII (Pre-type III collagen): reflecting the synthesis of type III collagen in the liver, the serum level is consistent with the degree of hepatic fibrosis and significantly correlates with the serum γ-globulin level. PCIII is closely related to the degree of activity of hepatic fibrosis formation. 2. HA (hyaluronidase): extremely elevated in liver cirrhosis. It can accurately and sensitively reflect the amount of fiber that has been generated in the liver and the damage to the hepatocytes.3. LN (laminin): a unique non-collagenous structural protein in the basement membrane, which can reflect the progress and severity of liver fibrosis.4. CIV (collagen type IV): the main component of the basement membrane, the increase in the content of which can reflect the process of hepatic fibrosis, and it is one of the early signs of hepatic fibrosis. These indicators are mainly used to diagnose the condition of chronic liver disease patients and the effect of treatment and are an important basis for measuring the activity of inflammation and the degree of fibrosis.

### Assessment of risk of bias

To assess the risk of bias, we will utilize tools from the Cochrane Collaboration [22].If any disagreements arise during this process, they will be resolved through consultation or third party review (JLZ).Risk of bias tool in V.5.1.0.Each included study was independently assessed by two researchers (XGL and SN).Based on the above tool, the risk of bias was ultimately categorized as low, high or unclear. Discrepancies, if any, will be resolved through discussion with the third author (JLZ).

### Measures of treatment effect

Data was analyzed using StataSE version 16. However, to account for heterogeneity that may not be explained among the selected trials, we applied a random effects meta-analysis to estimate all measures [23]. Publication bias was evaluated using Egger’s regression test when the outcomes were assessed with more than 10 trials. The levels of ALT, AST, GGT, TG, TC, PCIII, CIV, LN, and HA in each study were obtained through hospital tests, and the post-treatment test values for the test and control groups were directly selected as statistical parameters. Data for continuous variables were measured using the mean difference (MD) with a 95% confidence interval (CI).

### Level of evidence

The grading (level) of recommendations, assessments, developments and evaluations is used to assess the level of evidence for each outcome. The level of evidence was categorized as high, moderate, low or very low. Several aspects were considered in the assessment of the level of evidence, including risk of bias, imprecision, inconsistency, indirectness, publication bias, mass effect, dose-response and confounding. Criteria set by [24] Grade were used to assess these areas.

### Assessment of heterogeneity with reporting bias

Heterogeneity was assessed by calculating I^2^ using the Higgins test; an I^2^ value greater than 50% may indicate significant heterogeneity. Potential causes of heterogeneity were examined by sensitivity and subgroup analyses as follows:1) different combinations of electroacupuncture, and 2) number of weeks of electroacupuncture intervention. Funnel plots were used to assess publication bias if there were more than 10 eligible studies. Meanwhile, Egger’s [25] test was used to verify the presence of publication bias.

## Results

### Study characteristics

From the seven databases used in the initial search, 361 potentially relevant studies were identified (PubMed, n=51; Embase, n=27; Cochrane Library, n=9; Web of Science, n=3; WanFang, n=103; Cnki, n=101; VIP, n=67).After excluding duplicates, 165 articles were screened for inclusion in the full-text review based on title and abstract. Ultimately, 78 studies passed this stage. Subsequently,11 studies [26,27,28,29,30,31,32,33,34,35,36] were included after exclusion of 67 studies. The included studies focused on the most relevant transaminases in NAFLD as well as cholesterol and triglycerides. The PRISMA flowchart illustrates the study selection process (Figure 1). Table 1 displays the detailed characteristics of the studies.

**Figure 1.**
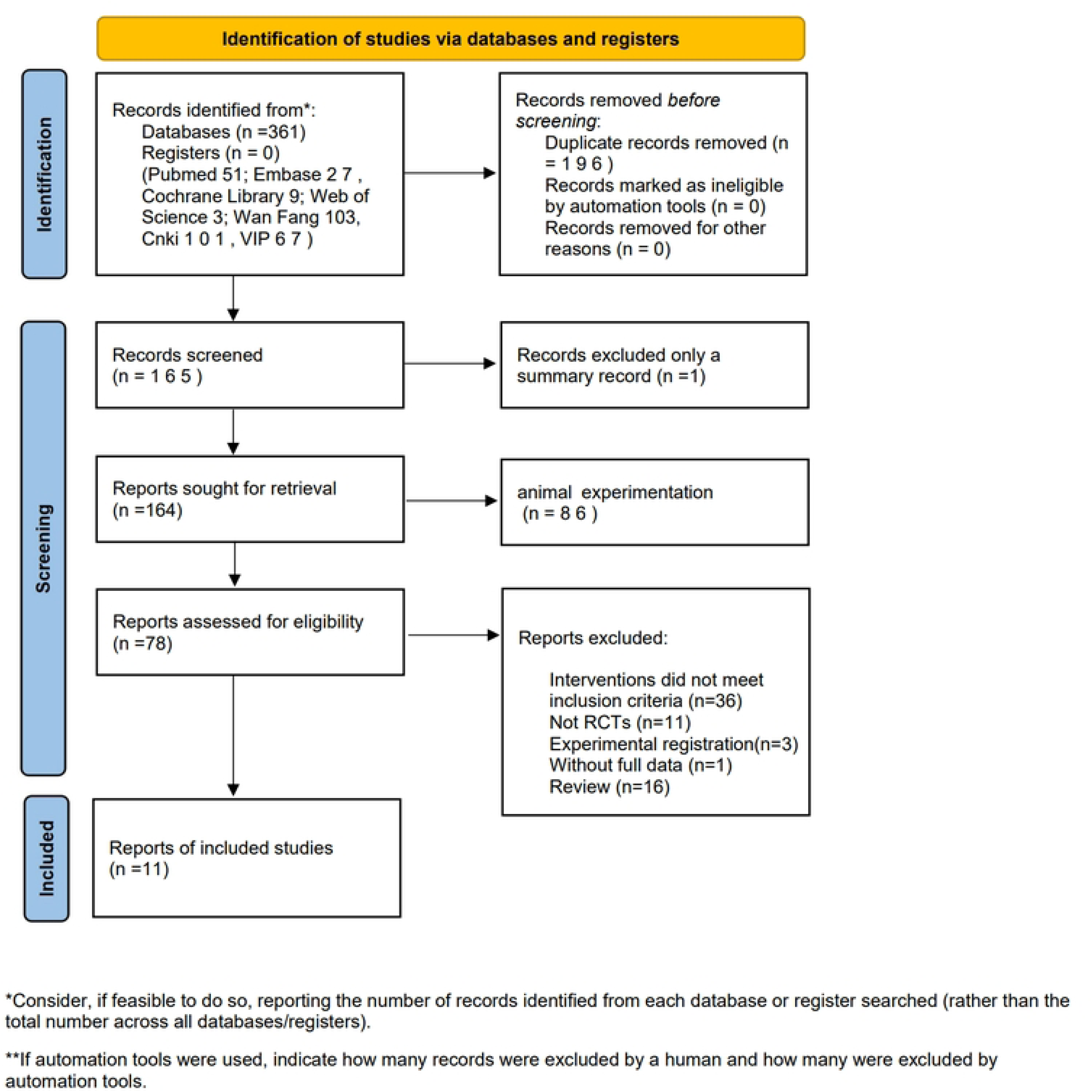
PRISMA 2020 flow diagram for new systematic reviews which included searches of databases and registers only. *From:* Page MJ, McKenzie JE, Bossuyt PM, Boutron I, Hoffmann TC, Mulrow CD, et al. The PRISMA 2020 statement: an updated guideline for reporting systematic reviews. BMJ 2021;372:n71. doi: 10.1136/bmj.n71

**Table 1:**
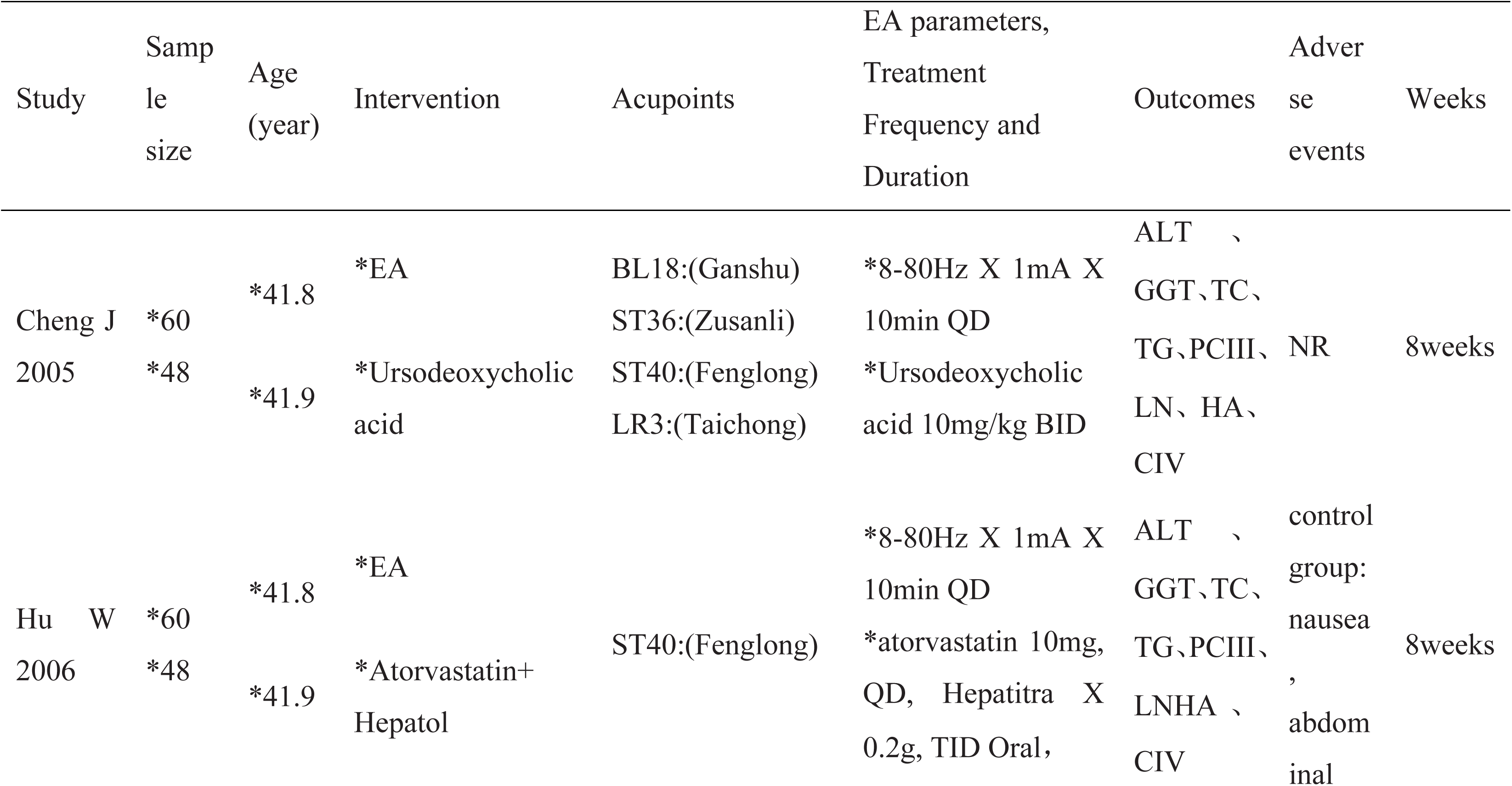
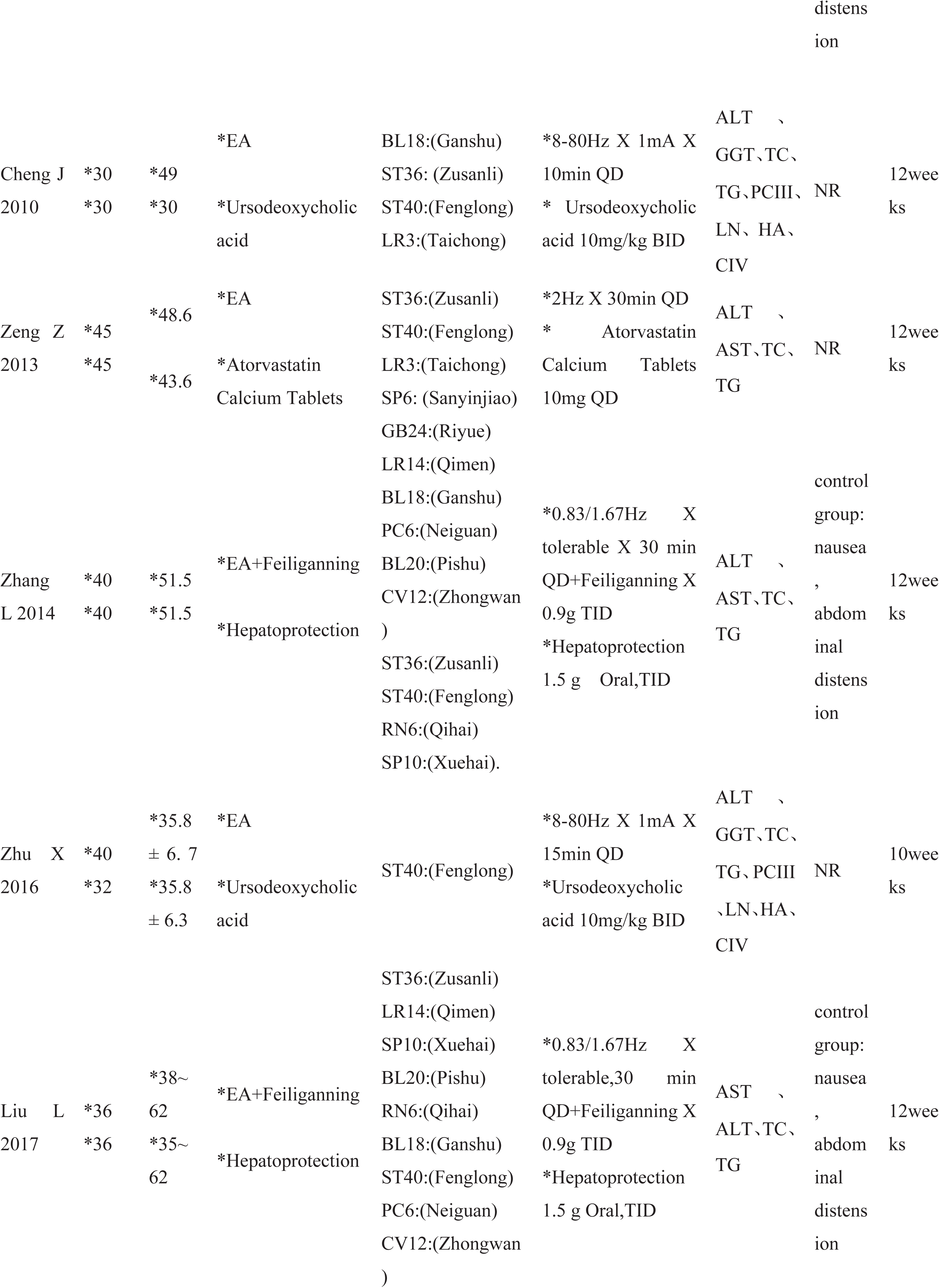
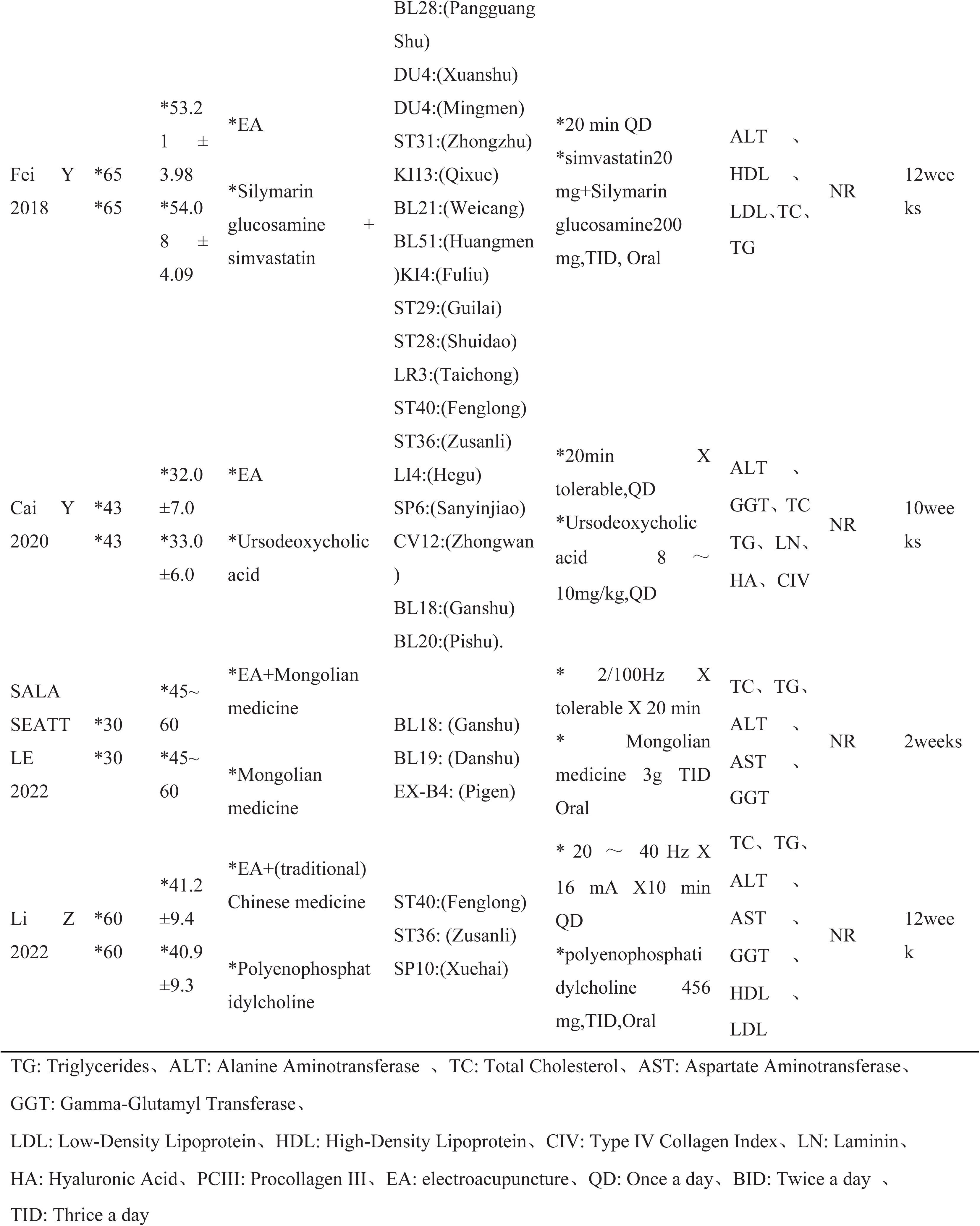
The detailed characteristics of the studies.

### Electroacupuncture therapy interventions

Electroacupuncture was added to the treatment of NAFLD in all 11 included studies. Eight studies [26,27,28,29,31,33,34,35] used electroacupuncture alone for treatment and three studies [30,32,36] used electroacupuncture in combination with medications.

All 11 studies compared the effects of electroacupuncture with the use of drugs alone. The experiments used a variety of acupoints for treatment, with the most frequently used being the Fenglong point, 9 times. The next most frequent was the Zusanli point, seven times. All these points had the effect of easing the liver and lowering lipids and regulating qi and blood. Five other studies used Ganshu. In 4 studies [26,27,28,37], electroacupuncture duration was 10 minutes. In 1 study [32], electroacupuncture duration was 15 minutes. In 3 studies [34,35,36], electroacupuncture duration was 20 minutes. In 3 studies [29,30,33], electroacupuncture lasted 30 minutes. Out of a total of 11 studies, the shortest duration of treatment was a 2-week study [36], and the longest duration of treatment was 12 weeks [28,29,30,32,33,36].

### Control interventions

Patients in 11 studies were treated with conventional western oral therapy to control the development of NAFLD. ursodeoxycholic acid was used in 4 studies [26,28,31,34] and hepatoprotection in 2 studies [30,32].Three of the studies used a combination of atorvastatin and hepatic Tylenol [27], a combination of different types of Montana [35], and a combination of simvastatin and silymarin glucosamine [33].There were also 2 studies without combinations, using Lipitor alone [29], and polyene phosphatidylcholine [36].

### Outcome measures

TG was reported in 11 studies [26,27,28,29,30,31,32,33,34,35,36], ALT in 11 studies [26,27,28,29,30,31,32,33,34,35,36], AST in 11 studies [26,27,28,29,30,31,32,33,34,35,36], and TC in 5 studies [29,30,32,35,36].TC, 5 studies [29,30,32,35,36] reported AST, 7 studies [26,27,28,31,34,35,36] reported GGT, 5 studies [26,27,28,31,34] reported CIV, 5 studies [26,27,28,31,34] reported LN, 5 studies [26,27.28,31,34] reported HA, and 4 studies [26,27,28,31] reported PCIII. Notably, only 3 studies [27,30,32] reported adverse events.

### Risk of bias

The quality of the studies included was assessed as low to moderate according to the Cochrane Risk of Bias (RoB) tool V.5.1.0. Four studies used a randomized grouping method, including one randomized lottery [34], and three used a randomized table of numbers method [27,31,36]. We considered these 4 studies to have a low risk of bias. Randomization methods were not specified in the remaining 7 studies. Given the specificity of electroacupuncture, blinding electroacupuncture implementers was not feasible. Therefore, we considered all 11 studies to have a high risk of performance bias. None of the studies identified any missing results or selective reporting, which we assessed as having a low risk of bias. Other biases were not explicitly discussed in any of the trials. Detailed results are shown in Table 2. The Risk of bias graph/summary (Supplemental 6).

**Table 2.** Cochrane ROB Table.

| Study | Random sequence generation (selection bias) | Allocation concealment (selection bias) | Blinding of participants and personnel (performance bias) | Blinding of outcome assessment (detection bias) | Incomplete outcome data (attrition bias) | Selective reporting (reporting bias) | Other bias |
| --- | --- | --- | --- | --- | --- | --- | --- |
| Cheng J 2005 | Unclear | Unclear | Unclear | Unclear | Low | Low | Unclear |
| Hu W 2006 | Low | Unclear | Unclear | Unclear | Low | Low | Unclear |
| Cheng J 2010 | Unclear | Unclear | Unclear | Unclear | Low | Low | Unclear |
| Zeng Z 2013 | Unclear | Unclear | Unclear | Unclear | Low | Low | Unclear |
| Zhang L 2014 | Unclear | Unclear | Unclear | Unclear | Low | Low | Unclear |
| Zhu X 2016 | Low | Unclear | Unclear | Unclear | Low | Low | Unclear |
| Liu L 2017 | Unclear | Unclear | Unclear | Unclear | Low | Low | Unclear |
| Fei Y 2018 | Unclear | Unclear | Unclear | Unclear | Low | Low | Unclear |
| Cai Y 2020 | Low | Unclear | Unclear | Unclear | Low | Low | Unclear |
| SALA SEATTLE 2022 | Unclear | Unclear | Unclear | Unclear | Low | Low | Unclear |
| Li Z 2022 | Low | Unclear | Unclear | Unclear | Low | Low | Unclear |

### Outcomes

#### TG(Triglycerides)

A total of 11 studies [26,27,28,29,30,31,32,33,34,35,36] were conducted on 986 patients with NAFLD. Of these, 509 were in the electroacupuncture group and 477 were in the drug control group. The heterogeneity test showed moderate statistical heterogeneity in the study. (P=0.025, I²=51.3%) (Supplemental 1 Figure 1). A random effects model was used. The results showed that there was a significant difference in TG between the electroacupuncture group and the control group (SMD=-0.84, 95%CI=-1.03, −0.65, P=0.000).

#### ALT (Alanine aminotransferase)

A total of 11 studies [26,27,28,29,30,31,32,33,34,35,36] were conducted on 986 patients with NAFLD. Of these, 509 were in the electroacupuncture group and 477 were in the drug control group. Heterogeneity test showed high statistical heterogeneity in the study.(P<0.000, I²=91.6%).Therefore, a random effects model was used in the meta-analysis revealing a significant difference between the electroacupuncture group and the conventional drug group in reducing ALT (SMD=-1.48, 95%CI=-1.98,-0.98 ,P=0.000) (Supplemental 1 Figure 2).After sensitivity analysis results showed that excluding the study of Fei [33], there were 856 patients in the remaining 10 studies, of which 444 were in the electroacupuncture group and 412 in the conventional drug group. The groups showed moderate heterogeneity (P<0.000, I²=52.2%) and the results showed statistically significant difference in ALT index between electroacupuncture group and conventional drug group (SMD=-1.18, 95%CI=-1.39, −0.97, P=0.000).

#### TC (Total cholesterol)

A total of 11 studies [26,27,28,29,30,31,32,33,34,35,36] were conducted on 986 patients with NAFLD. Of these, 509 were in the electroacupuncture group and 477 were in the drug control group. Heterogeneity test showed high statistical heterogeneity in the study (P=0.002, I²=64.0%). The results of the study showed that there was a significant difference between the effect of electroacupuncture and drug groups on total cholesterol. (SMD=-0.59, 95%CI=-0.81, −0.38, P=0.000) (Supplemental 1 Figure 3). The results of sensitivity analysis showed that excluding the study of Li [36], there were 866 patients in the remaining 10 studies. Among them, 449 were in the electroacupuncture group and 417 in the control group. The groups did not show heterogeneity (P=0.894, I²=0%). Therefore, we performed the analysis again. The results of meta-analysis showed that there was a significant difference in TC effect between the electroacupuncture group and the control group (SMD=-0.48, 95%CI=-0.62, −0.35, P=0.000). Consistency was observed between the results.

#### GGT (Gamma-glutamyl transpeptidase)

A total of 7 studies [26,27,28,31,34,35,36] were conducted on 614 patients with NAFLD. Of these, 323 patients were in the electroacupuncture group, and 291 patients were in the control group. The heterogeneity test showed high statistical heterogeneity in the study. (p<0.000, I²=77.6%). Analyzing the data using random effects model, the results of the study showed that there was a significant difference in the effect of electroacupuncture group and control group on gamma-glutamyl transpeptidase. (SMD=-1.21, 95%CI=-1.59, −0.84, P=0.000) (Supplemental 1 Figure 4). The results of the sensitivity analysis showed that excluding the study of SALA SEATTLE [35], there were 554 patients in the remaining 6 studies. Of these, 293 were in the electroacupuncture group and 261 in the control group. The groups showed moderate to low heterogeneity (P=0.234, I²=26.7%). We still used a random effects model, and the meta-analysis showed a significant difference in the GGT effect between the electro-acupuncture group and the control group (SMD=-1.36, 95%CI=-1.58, −1.14, P=0.000).

#### AST (Aspartate aminotransferase)

A total of five studies [29,30,32,35,36] were conducted on 422 patients with NAFLD. 211 patients were in the electroacupuncture group, and 211 patients were in the control group. The heterogeneity test showed low heterogeneity in the study. (P=0.234, I²=28.2%). A random effects model was used to analyze the data. The results of the study showed that there was a significant difference between the effect of electroacupuncture group and control group on AST aspartate aminotransferase. (SMD=-0.84, 95%CI=-1.08, −0.60, P=0.000) (Supplemental 1 Figure 5). The results of sensitivity analysis showed that excluding the study by Zeng 2013, there were 332 patients in the remaining four studies. Of these, 166 were in the electroacupuncture group and 166 in the control group. The groups did not show heterogeneity (P=0.750, I²=0%). Meta-analysis results showed a significant difference in AST effect between electroacupuncture and control groups (SMD=-0.95, 95%CI=-1.18, −0.72, P=0.000). Consistency was observed between the results.

#### CIV (Collagen type IV)

A total of 5 studies [26,27,28,31,34] were conducted on 434 patients with NAFLD. Of these, 233 patients were in the electroacupuncture group, and 201 patients were in the control group. The heterogeneity test showed that there was no statistical heterogeneity in the study. (P=0.965, I²=0%). The results of the study showed that there was a significant difference in the effect of electroacupuncture group and control group on type IV collagen. (SMD=-1.08, 95%CI=-1.28, −0.87, P=0.000) (Supplemental 1 Figure 6).

#### LN (Laminin)

A total of 5 studies [26,27,28,31,34] were conducted on 434 patients with NAFLD. Of these, 233 patients were in the electroacupuncture group, and 201 patients were in the control group. The Heterogeneity test showed that there was no statistical heterogeneity in the study. (P=1.000, I²=0%). The results of the study showed that there was a significant difference in the effect of electroacupuncture group and control group on laminin. (SMD=-1.90, 95%CI=-2.13, −1.67, P=0.000) (Supplemental 1 Figure 7).

#### HA (Hyaluronidase)

A total of 5 studies [26,27,28,31,34] were conducted on 434 patients with NAFLD. Of these, 233 patients were in the electroacupuncture group, and 201 patients were in the control group. The heterogeneity test showed that there was no statistical heterogeneity in the study. (P=1.000, I²=0%). The results of the study showed that there was a significant difference in the effect of electroacupuncture group and control group on type IV collagen. (SMD=-3.42, 95%CI=-3.72, −3.12, P=0.000) (Supplemental 1 Figure 8).

#### PCIII (Pre-type III collagen)

A total of four studies [26,27,28,31] were conducted on 348 patients with NAFLD. Of these, 190 patients were in the electroacupuncture group, and 158 patients were in the control group. The heterogeneity test showed that there was no statistical heterogeneity in the study. (P=1.000, I²=0%). The results of the study showed that there was a significant difference in the effect of electroacupuncture group and control group on type III collagen. (SMD=-0.41, 95%CI=-0.62, −0.19, P=0.000) (Supplemental 1 Figure 9).

### Adverse events

Adverse events were reported in only three studies, including [27, 30, 32]. The adverse events that occurred in all three studies were nausea and abdominal distension in some patients within the western medicine treatment group. A meta-analysis was not conducted because the number of adverse reactions in the western drug group could not be determined.

### Subgroup analysis

We performed subgroup analyses of the metrics: A) different combinations of electroacupuncture. B) number of weeks of electroacupuncture intervention. The results are detailed in Table 3.

**Table 3.**
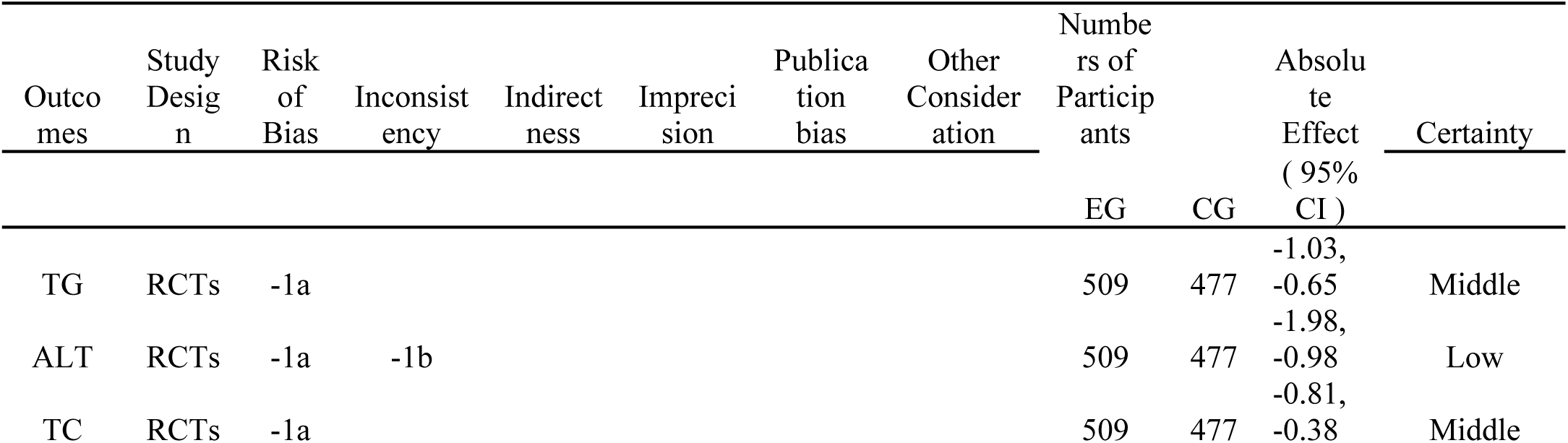
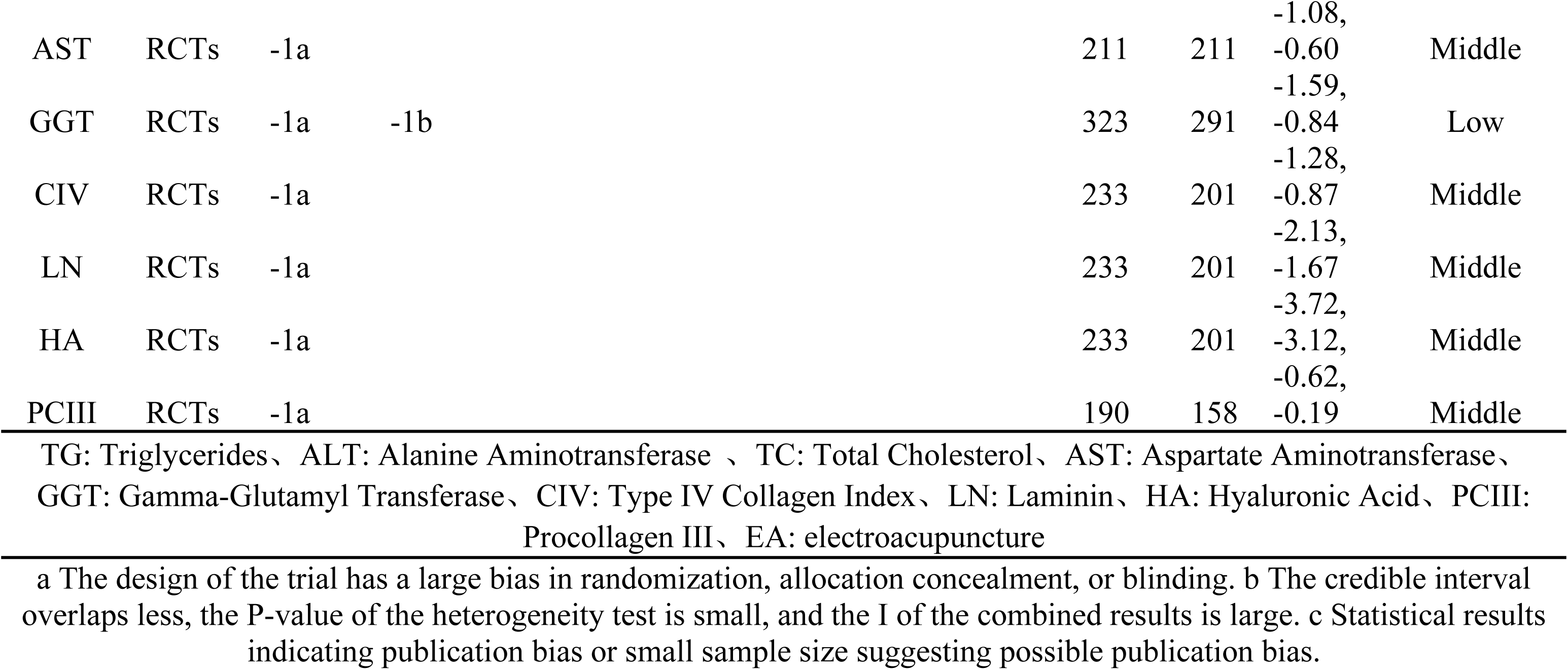
Quality of evidence.

#### Subgroup analysis of TG

##### Different combinations of electroacupuncture

In the subgroup analysis of electroacupuncture alone, low heterogeneity was found between 8 studies (I²=26.5%), which showed a significant difference between the electroacupuncture group and the control group in terms of lowering triglycerides (SMD=-0.89, 95%CI=-1.017, −0.71, P=0.000). In the subgroup analysis of electroacupuncture combined with drugs, a high degree of heterogeneity was found between the three studies (I²=79.9%). Analyses using a random-effects model showed a significant difference between electroacupuncture combined with drugs and controls in reducing TG (SMD=-0.69, 95%CI=-1.25, −0.13, P=0.015). (Supplemental 2 Figure 1)

##### Different weeks of intervention

In the subgroup analysis of weeks of intervention not greater than 8 weeks, no heterogeneity was found in 3 studies (I²=0%). The analyses showed a significant difference between the electroacupuncture group and the control group in terms of TG reduction (SMD=-0.93, 95%CI=-1.18, −0.68, P=0.000). In the subgroup analysis of weeks of intervention greater than 8 weeks, the test for heterogeneity showed that in 8 studies (I²=64.9%). Analyses using a random-effects model showed a significant difference between the electroacupuncture and control groups in reducing TG (SMD=-0.80, 95% CI=-1.07, −0.54, P=0.000). (Supplemental 2 Figure 2)

#### Subgroup analysis of ALT

##### Different combinations of electroacupuncture

In the subgroup analysis of electroacupuncture alone, 8 studies (I²=93.8%) found high heterogeneity. Random effects model analysis showed a significant difference between electroacupuncture alone and control in reducing ALT (SMD=-1.44, 95%CI=-2.12, −0.76, P=0.000). In the subgroup analysis of electroacupuncture combined with drugs, no significant heterogeneity was found between the 3 studies (I²=0). The analyses showed a significant difference between electroacupuncture combined with drugs and controls in reducing ALT (SMD=-1.58, 95%CI=-1.86, −1.31, P=0.000). (Supplemental 2 Figure 3)

##### Different weeks of intervention

In the subgroup analysis of weeks of intervention not greater than 8 weeks, no heterogeneity was observed in the 3 studies (I²=0%). Analyses using a random-effects model showed a significant difference between the electroacupuncture and control groups in reducing ALT (SMD=-1.02, 95% CI=-1.27, −0.77, P=0.000).In the subgroup analysis where the number of weeks of intervention was greater than 8 weeks, the heterogeneity test showed that in 8 studies (I²=93.6%).Analyses using a random effects model showed a significant difference between the electroacupuncture group and the control group in reducing ALT (SMD=-1.67, 95% CI=-2.36, −0.98, P=0.000). (Supplemental 2 Figure 4)

#### Subgroup analysis of TC

##### Different combinations of electroacupuncture

In the subgroup analysis of electroacupuncture alone, no heterogeneity was observed in 8 studies (I²=0%). Modelling analysis showed a significant difference between electroacupuncture alone and the control group in terms of TC reduction (SMD=-0.45, 95%CI=-0.59, −0.30, P=0.000). In the subgroup analysis of electroacupuncture combined with drugs, a high degree of heterogeneity was found between the three studies (I²=82.1%). Random effects model analysis showed a significant difference between electroacupuncture combined with drugs and control in reducing TC (SMD=-0.96, 95%CI=-1.57, −0.36, P=0.002). (Supplemental 2 Figure 5)

##### Different weeks of intervention

In the subgroup analysis of weeks of intervention not greater than 8 weeks, no heterogeneity was observed in the 3 studies (I²=0%). Analyses using a random-effects model showed no significant difference between the electroacupuncture and control groups in terms of TC reduction (SMD=-0.47, 95% CI=-0.71, −0.23, P=0.000). In the subgroup analysis of weeks of intervention greater than 8 weeks, high heterogeneity was found in 8 studies (I²=73.5%). Analyses using a random-effects model showed a significant difference between the electroacupuncture and control groups in terms of TC reduction (SMD=-0.65, 95%CI=-0.94, −0.35, P=0.000). (Supplemental 2 Figure 6)

#### Subgroup analysis of AST

##### Different combinations of electroacupuncture

In the subgroup analysis of electroacupuncture alone, low heterogeneity was observed in 2 studies (I²=6.9%). Effect model analysis showed a significant difference between electroacupuncture alone and control in reducing AST (SMD=-0.58, 95% CI=-0.92, −0.24, P=0.001). In the subgroup analysis of electroacupuncture combined with drugs, no heterogeneity was observed between the 3 studies (I²=0%). There was a significant difference between electroacupuncture combined with drugs and controls in reducing AST (SMD=-0.99, 95%CI=-1.24, −0.73, P=0.000). (Supplemental 2 Figure 7)

#### Subgroup analysis of GGT

##### Different combinations of electroacupuncture

In the subgroup analysis of electroacupuncture alone, high heterogeneity was observed in six studies (I²=80.3%). Random effects model analysis showed a significant difference between electroacupuncture alone and control in reducing GGT (SMD=-1.25, 95% CI=-1.70, −0.81, P=0.000). (Supplemental 2 Figure 9)

##### Different weeks of intervention

In the subgroup analysis of weeks of intervention not greater than 8 weeks, a high degree of heterogeneity was found in 3 studies (I²=88.0%). Analyses using random effects model showed a significant difference between the electroacupuncture group and the control group in terms of GGT reduction (SMD=-0.95, 95%CI=-1.69, −0.20, P=0.013). In the subgroup analysis where the number of weeks of intervention was >8 weeks, moderate heterogeneity was observed in 4 studies (I²=56.0%). Analyses using a random-effects model showed a significant difference between the electroacupuncture and control groups in reducing GGT (SMD=-1.40, 95% CI=-1.77, −1.03, P=0.000). (Supplemental 2 Figure 10)

### Sensitivity analyses

Sensitivity analyses of the data were performed using Stata V16.0 software. The results showed that the sensitivity of the data in each group was basically stable.

The results of sensitivity analysis of ALT level showed that Fei [33] was the main source of heterogeneity. The results of sensitivity analysis of AST level showed that Zeng [29] was the main source of heterogeneity. The results of sensitivity analysis at the TC level indicated that Li [36] was the main source of heterogeneity. The results of sensitivity analyses at the GGT level indicated that SALA SEATTLE [35] was the main source of heterogeneity. (Supplemental 3)

### Publication bias

Publication bias was visually analyzed using funnel plots for the 3 included studies with more than 10 main indicators, all of which were subjected to the egger test. The funnel plot of TG showed a slight asymmetry, suggesting some publication bias. egger test, p=0. 493. The funnel plot of ALT showed a slight asymmetry, suggesting some publication bias. egger test, p=0.028. The funnel plot of TC showed a slight asymmetry, suggesting some publication bias. egger test, p=0.732. None of them had significant publication bias. Combined with the results of sensitivity analyses, it was hypothesized that it might be caused by heterogeneity due to some of the studies. (Supplemental 4)

### GRADE evaluation

Five downgrading factors (i.e., risk of bias, inconsistency, indirectness, imprecision, and publication bias) were considered in the quality of evidence assessment of this study. Of all 11 studies, 4 mentioned the randomization allocation method and 7 did not specify the randomization method. There was no blinding in the 11 studies, so the risk of bias was downgraded by 1 level for all studies. For studies of ALT, GGT, there was a high degree of between-study heterogeneity, so inconsistency was downgraded.TC studies had a high degree of between-study heterogeneity, but we identified sources of heterogeneity and did not reduce it. Overall, the level of evidence was moderate for TG, TC, CIV, LN, HA, PCIII, and AST and low for ALT and GGT. Detailed results are shown in Table 4.

**Table 4.**
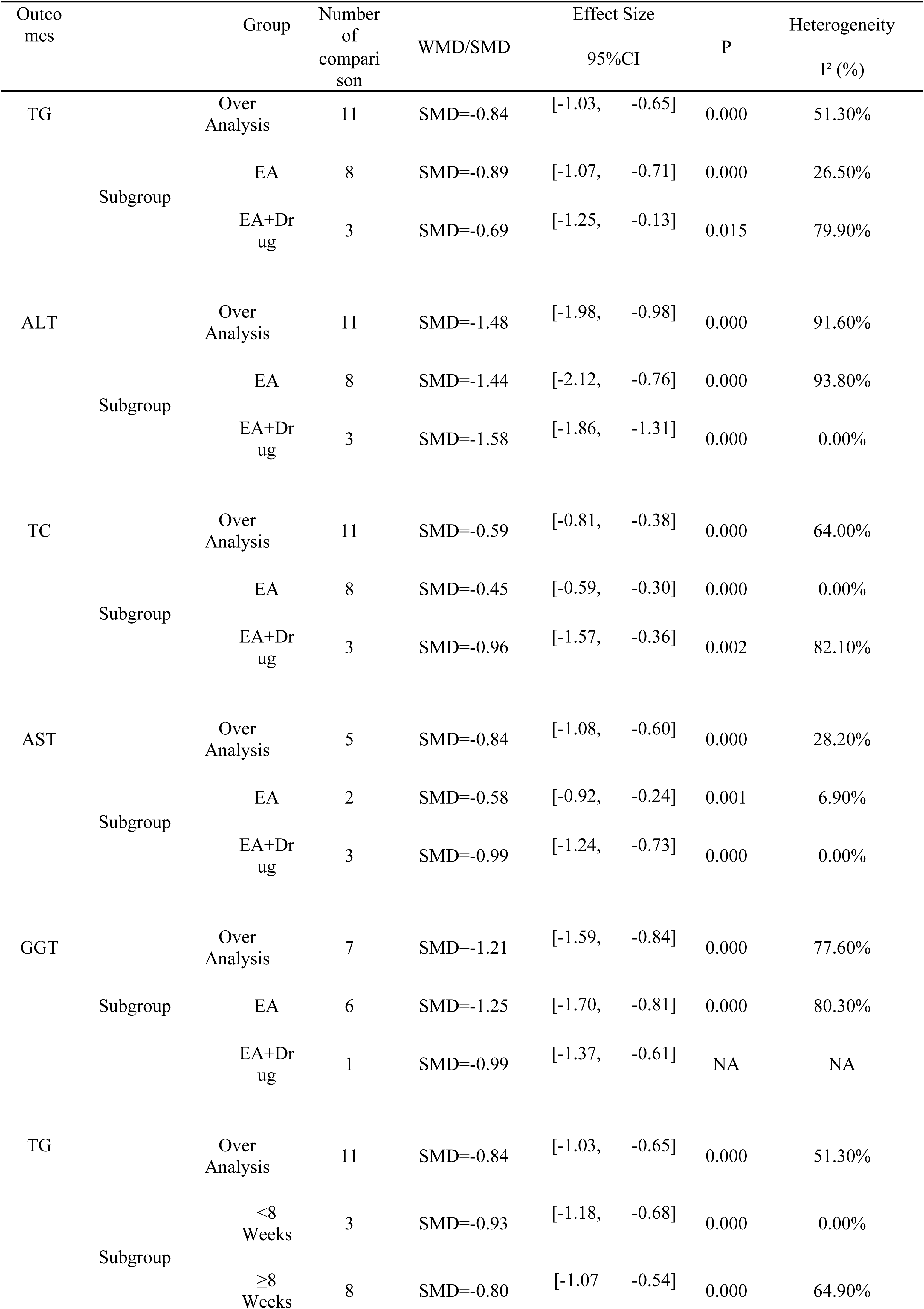
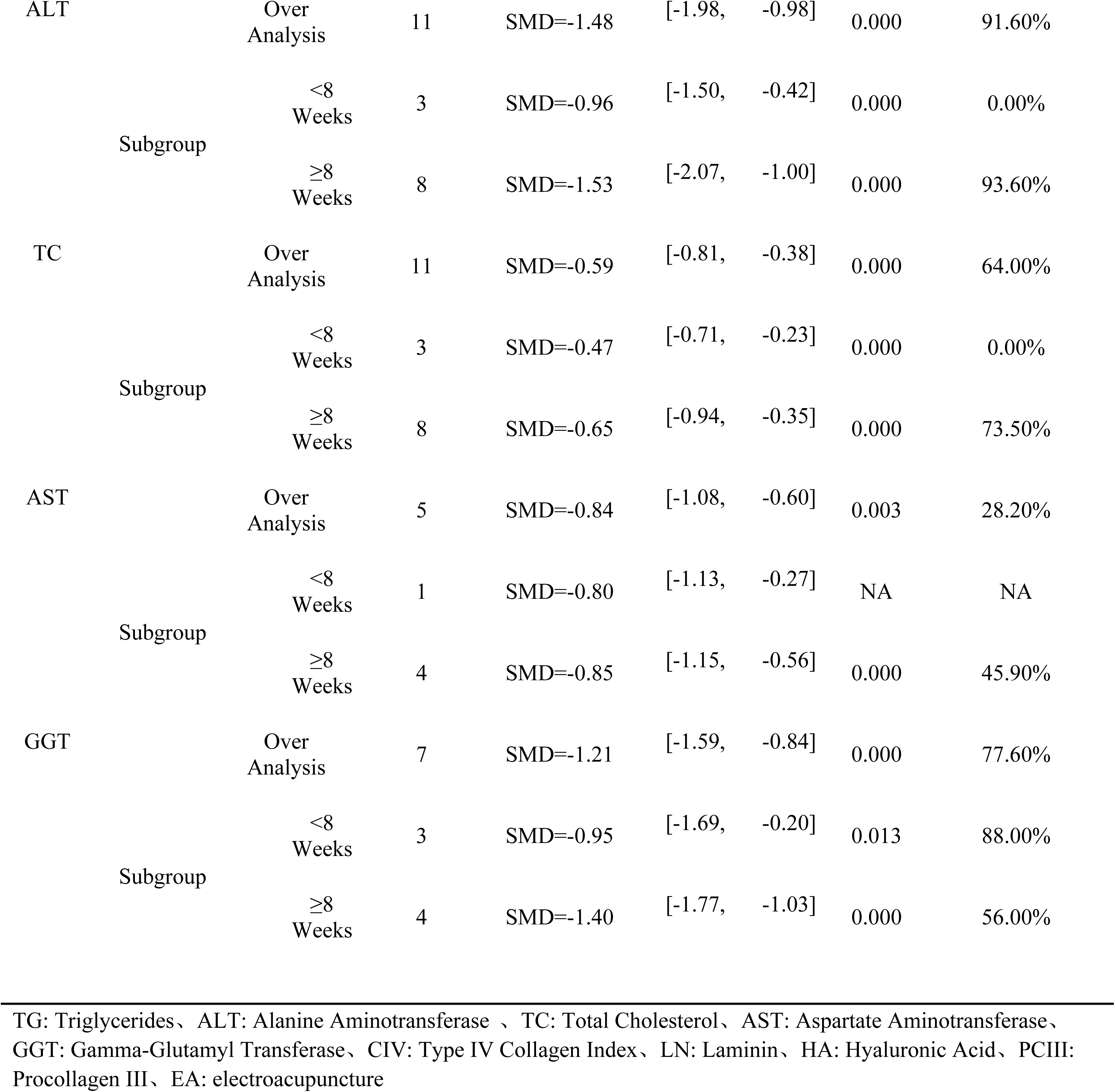
Results of the analysis of individual outcome indicators and their subgroups.

## Discussion

### Main results

This systematic review examined 11 randomized controlled trials investigating electroacupuncture for NAFLD. To our knowledge, this is the first systematic review assessing electroacupuncture for NAFLD and follows the preferred reporting items for systematic reviews and meta-analyses.

Cirrhosis and hepatocellular carcinoma represent the most severe outcomes of disease progression in patients with non-alcoholic fatty liver disease (NAFLD). The current meta-analysis indicates that acupuncture and traditional treatments exhibit comparable effectiveness and safety in addressing the early symptoms of NAFLD. Electroacupuncture, either alone or in combination with drugs, had a significant effect on improving ALT, AST, TC, TG and LN, HA, PC III, C IV levels and halting the progression of fibrosis compared to drugs alone.

After summarizing the electroacupuncture stimulation methods utilized in the study, the results indicated that the designated stimulation duration ranged from 10 to 30 minutes. Regarding the selection of acupoints for electroacupuncture stimulation, the Fenglong acupoint (ST40), located on the anterolateral side of the calf, is found 8 inches above the tip of the lateral ankle, outside the Strip Mouth acupoint, and two transverse (middle) fingers from the anterior edge of the tibia. The Zusanli acupoint (ST36), located on the anterolateral side of the calf, one transverse finger away from the anterior edge of the tibia, were the most frequently used points for needle stimulation. Overall, the application of electroacupuncture stimulation was primarily concentrated on the anterolateral and anteromedial aspects of the calf. It is important to note that the intensity of electroacupuncture stimulation is clinically adjusted according to the patient’s tolerance level.

A study published by Peiwen Chen [37] aligns with our findings. The meta-analysis demonstrated that the use of electroacupuncture significantly improved both transaminase levels and lipid-lowering effects in patients with non-alcoholic fatty liver disease (NAFLD). Additionally, the effects on liver fibrosis markers indicated that electroacupuncture could halt the progression of liver fibrosis in these patients. Therefore, it can be concluded that electroacupuncture is a highly feasible complementary alternative therapy for patients who are unable to use systemic medications for long-term lipid reduction and liver protection due to underlying health conditions or contraindications.

In our meta-analysis, there was a great heterogeneity in the studies on ALT (I²=91.6%) and GGT (I²=77.6%) in patients with NAFLD, and a moderate heterogeneity in the studies on AST (I²=28.2%), TG (I²=51.3%) and TC (I²=64.0%) in patients with NAFLD. We performed subgroup analyses, which showed a substantial decrease in heterogeneity in the studies of electroacupuncture alone (I²=6.9%) and electroacupuncture combined with drugs (I²=0) in the subgroup analyses of AST, which led to the conclusion that the mode of intervention was the source of heterogeneity in AST. Other subgroup analyses did not show a significant source of heterogeneity. In addition, we identified a source of heterogeneity in studies of ALT levels by sensitivity analysis (P=0.000, I²=91.6%) Fei [33] was the main source of heterogeneity. After the exclusion of Fei[33], the heterogeneity between studies was moderate (P=0.000, I²=52%).We identified Zeng [29] as a source of heterogeneity in the AST level studies (P=0.000, I²=28.2%), and the heterogeneity between studies disappeared after the exclusion of the study by Zeng 2013 [29] (P=0.000, I²=0%).We identified Li [36] as the source of heterogeneity in the TC level studies (P=0.000, I²=64%), and the heterogeneity between studies disappeared after excluding the study of Li [36] (P=0.000, I²=0%) For this we hypothesized that the source of heterogeneity might be related to different drug combinations.

Current research indicates that the mechanisms underlying disease pathogenesis and progression in non-alcoholic fatty liver disease (NAFLD) are diverse and complex. The progression of NAFLD is a multifaceted process involving the interaction of various mechanisms. Insulin resistance is a central factor that increases lipid synthesis, promotes the transport of fatty acids to the liver, and inhibits their β-oxidation, ultimately leading to hepatic lipid accumulation. Genetic factors, particularly polymorphisms in genes such as PNPLA3, TM6SF2, and GCKR, are associated with the onset and severity of NAFLD, influencing hepatic lipid content and inflammatory responses. Oxidative stress and endoplasmic reticulum stress result in an increase in reactive oxygen species, which activate inflammation and hepatic stellate cells, thereby promoting liver fibrosis. Biotoxicity, particularly from free cholesterol, exacerbates sterile inflammation by interacting with the YAP-TAZ pathway. Dysbiosis of the intestinal microbiota disrupts the intestinal mucosal barrier, leading to increased endotoxins and inflammatory factors that trigger hepatic inflammation and fibrosis. Collectively, these mechanisms drive the progression of NAFLD from mild to severe fatty liver [38,39]. Our meta-analysis revealed significant changes in multiple biomarkers of disease in response to electroacupuncture. The progression of non-alcoholic fatty liver disease (NAFLD) is closely linked to alterations in various biomarkers. These include elevated levels of liver enzymes such as alanine aminotransferase (ALT), aspartate aminotransferase (AST), and γ-glutamyl transferase (GGT), which typically indicate hepatocellular damage. Additionally, increased levels of triglycerides (TG) and total cholesterol (TC) reflect abnormalities in lipid metabolism. Furthermore, elevated indicators of liver fibrosis, such as procollagen type III (PCIII), hyaluronidase (HA), laminin (LN), and collagen type IV (CIV), suggest the formation and progression of hepatic tissue fibrosis. These biomarkers are crucial for assessing the severity of NAFLD and the effectiveness of treatment.

In the treatment of patients with non-alcoholic fatty liver disease (NAFLD), the effects are primarily achieved through long-term lipid-lowering strategies and liver protection. Several key acupoints included in this meta-analysis, such as Fenglong, Gan Shu, Zusanli, and Taichong, align with the acupoint selection used in current animal experiments [40,41]. Electroacupuncture at the Fenglong and Zusanli points demonstrated a beneficial regulatory effect on rats with non-alcoholic fatty liver disease. The proposed mechanism of action may involve the downregulation of the SREBP-1c gene and protein expression, improvement of endoplasmic reticulum stress, and regulation of lipid metabolism disorders. Consequently, this treatment reduces inflammatory damage to liver tissues, with electroacupuncture at the Fenglong point showing superior effects [40]. Electroacupuncture can inhibit hepatocyte cell death in rats with non-alcoholic fatty liver disease (NAFLD). Its mechanism of action may be associated with a reduction in serum lipopolysaccharide (LPS) levels and the downregulation of factors related to the non-classical pathway, including GSDMD, GSDMD-N, Caspase-11, IL-1β, IL-18, and TNF-α [41]. Additionally, the study controlled for other biomarkers and similarly confirmed the significant effectiveness of electroacupuncture in NAFLD patients, particularly regarding the mechanisms of lipid formation and the generation of hepatic inflammation.

The included studies demonstrated a moderate risk of bias and heterogeneity in the meta-analysis, which typically results in a low level of evidence. Based on the graded evidence available in this study, we can only provide a weak recommendation for the use of electroacupuncture in treating non-alcoholic fatty liver disease (NAFLD). Nevertheless, our findings remain positive regarding the efficacy of electroacupuncture for NAFLD treatment. Future randomized controlled trials should consider more rationally designed electroacupuncture protocols, as these techniques allow for partial standardization. This approach would facilitate the acquisition of high-quality, evidence-based results. In conclusion, the mechanisms by which electroacupuncture affects the treatment of NAFLD are not fully understood, and further research is warranted. Given the slightly low level of evidence in this study, the results should be interpreted with caution. Ultimately, further validation of our findings will necessitate larger sample sizes and better-designed randomized controlled trials.

## Limitations

Firstly, electroacupuncture, as a distinctive form of complementary and alternative medicine, poses challenges in implementing blinding in research studies. Additionally, establishing precise specifications for electroacupuncture, particularly concerning stimulation intensity and point selection, remains difficult. Consequently, the effectiveness of electroacupuncture interventions may be influenced by bias.

Second, according to the NAFLD guidelines, drug selection should be based on the type and extent of liver damage, as well as the patient’s metabolic profile. It is recommended that different drug therapies be chosen based on the underlying disease conditions and the degree of liver injury. However, patients with coexisting cardiovascular disease or metabolic disorders were not included in our study analysis.

Finally, the NAFLD syndrome encompasses various degrees of progression, including NAFLD, NAFLD fibrosis, and cirrhosis. More clinical trials and further refined discussions are necessary to address the treatment of these different stages.

Previously, electroacupuncture has gained popularity as a treatment for various types of pain, but it also demonstrates significant efficacy in the management and treatment of chronic metabolic diseases. The results of our meta-analysis indicate that electroacupuncture is effective in treating patients with non-alcoholic fatty liver disease (NAFLD). Furthermore, electroacupuncture combined with pharmacological interventions proved to be more effective than drug therapy alone in reducing lipid levels and preventing the progression of liver fibrosis. These findings will provide clinicians with additional treatment options. We also recommend that patients consider electroacupuncture as a viable treatment for NAFLD. In conclusion, the underlying mechanisms by which electroacupuncture benefits patients with NAFLD require further investigation, and additional studies are necessary to clarify these mechanisms. Given the moderate quality of the included studies, the results should be interpreted with caution.

## Conclusion

The results of the meta-analysis suggest that electroacupuncture offers significant advantages in the treatment of non-alcoholic fatty liver disease (NAFLD). Furthermore, electroacupuncture combined with conventional drug therapy has been shown to reduce lipolysis and slow the progression of hepatic fibrosis compared to drug therapy alone more effectively. Based on the current evidence, we recommend using electroacupuncture therapy with the primary acupoints of Ganshu, Fenglong, Taichong, and Zusanli for the treatment of NAFLD. It is essential to conduct additional well-designed multicenter clinical trials with larger sample sizes and combination experiments for various metabolic diseases to validate and strengthen these findings.

## Abbreviations

NAFLD: Nonalcoholic Fatty Liver Disease
RCTs: Randomized Controlled Trials
WMD: Weighted Mean Difference
CI: Confidence Interval
SMD: Standard Mean Difference
PRISMA: Preferred Reporting Items for Systematic Reviews and Meta-Analyses
GRADE: Grading of Recommendations, Assessment, Development, and Evaluation
EG: Experimental Group
CG: Control Group
NI: No Intervention
NA: Not Applicable
EA: Electroacupuncture
TG: Triglycerides
TC: Total cholesterol
ALT: Alanine aminotransferase
AST: Aspartate aminotransferase
GGT: Gamma-glutamyl transpeptidase
LN: Laminin
HA: Hyaluronidase
PCIII: Pre-type III collagen
CIV: Collagen type IV

## Publisher’s note

All claims expressed in this article are solely those of the authors and do not necessarily represent those of their affiliated organizations, or those of the publisher, the editors and the reviewers. Any product that may be evaluated in this article, or claim that may be made by its manufacturer, is not guaranteed or endorsed by the publisher.

## Ethics and dissemination

The study did not involve participants’ personal data and therefore did not require ethical approval. This systematic review will be published in a peer-reviewed journal.

## Credits authorship contribution statement

XGL: Formal analysis, Investigation, Software, Writing – original draft, Writing – review & editing, Conceptualization, Data curation, Methodology. SN: Methodology, Writing – review & editing, Conceptualization, Formal analysis, Investigation. JLZ: Formal analysis, Software, Writing – review & editing, Data curation. YTW: Data curation, Writing – review & editing. LWZ: Funding acquisition, Project administration, Supervision, Writing – review & editing.

## Patient and public involvement

Patients and/or the public were not involved in the design, conduct, or reporting, or dissemination plans of this research.

## Patient consent for publication

Not applicable.

## Data Availability

All relevant data are within the manuscript and its Supporting Information files.

